# Modeling spontaneous hepatitis C virus clearance by age, sex, *IFNL4* genotype, and infection history: Implications for simulations of elimination and vaccine trials

**DOI:** 10.64898/2026.11.18.26368407

**Authors:** Keshav R. Gandhi, Sophia Mao, Eric Tatara, Jonathan Ozik, Basmattee Boodram, Scott J. Cotler, Marian Major, Alexander Gutfraind, Harel Dahari

**Author notes:** Corresponding authors (K.G.) & (H.D.).

## Abstract

**Background and Aims:** The probability of hepatitis C virus (HCV) clearance is mediated by an interaction between numerous factors. However, variation in spontaneous HCV clearance was previously modeled using relatively simple approaches with fixed probabilities by sex. Here, we developed a literature-informed clearance algorithm that considers age, sex, interferon lambda 4 (*IFNL4*) genotype, and infection history to model the probability of HCV clearance in vulnerable populations, such as people who inject drugs (PWID), and in controlled human infection (CHI) models for future HCV vaccines.

**Methods:** We modeled HCV clearance probabilities for three infection-history pathways: first infection, reinfection after spontaneous clearance, and reinfection after direct-acting antiviral (DAA) cure. Logistic and cumulative-hazard equations assigned person-specific clearance probabilities in the first two pathways. The post-DAA pathway used fixed clearance probabilities. We tested the algorithm using HepCEP (Hepatitis C Elimination in PWID), an agent-based simulator.

**Results:** First-infection clearance probabilities ranged from 0.13 to 0.51 across eight age, sex, and *IFNL4* genotype subgroups. Clearance probabilities for reinfection after spontaneous clearance ranged from 0.20 to 0.80 at literature point estimates. Across 100 runs, the mean assigned clearance probabilities were 0.27 for first infections and 0.52 for reinfections after spontaneous clearance, compared with literature inputs of 0.26 and 0.52, respectively. Probabilities varied across subgroups and runs.

**Conclusion:** Simulations reproduced the algorithm’s intended clearance heterogeneity and uncertainty by age, sex, *IFNL4* genotype, and infection history, which may support models of HCV elimination in vulnerable populations and CHI model development for future HCV vaccines.

## 1. Introduction

Despite the increasing availability of curative therapy, hepatitis C virus (HCV) infection remains a major public health burden in the United States, affecting an estimated 2.5–4 million adults with current infection in 2017–2020.^1^ Consequently, developing an effective HCV vaccine has become a prominent research priority.^2, 3^ Agent-based models (ABMs), which represent individual people and their interactions, have been used to simulate vaccine trials and characterize HCV transmission dynamics.^4, 5^ Simulated trials can assess study designs that would be difficult and expensive to test repeatedly in real life. Elimination models in people who inject drugs (PWID) can further estimate the treatment intensity and costs needed to reduce chronic infections.^5, 6^ In both applications, spontaneous clearance affects the proportion of infections that become chronic, influencing projected trial outcomes and treatment needs.

Spontaneous clearance of primary infections varies with the characteristics of the infected person. Pooled estimates of spontaneous clearance were 27.9% by 6 months and 37.1% by 24 months after infection.^7^ Factors associated with clearance of primary infection include age, sex, HCV genotype, and variants in the host *IFNL4* region, historically described as variants near interleukin 28B (*IL28B*, now *IFNL3*), a separate neighboring gene.^7–9^ Younger adults and people of female sex are more likely to clear a first infection.^7^ Variants in *IFNL4* can affect interferon- mediated antiviral responses,^10^ and their frequencies differ across racial and ethnic populations.^11–14^ Representing *IFNL4* genotype allows relevant population variation to be included without treating race or ethnicity itself as a biological cause of clearance. In particular, the CC genotype of rs12979860 is associated with a higher probability of spontaneous clearance than CT or TT.^8, 15^

Additionally, people who spontaneously clear an initial HCV infection may clear a later infection more readily and rapidly, with reported clearance proportions of approximately 50%–83% across studies of reinfection.^16–18^ Evidence for factors associated with clearance of reinfection is more limited. In the InC^3^ study, consistent with characteristics associated with clearance of primary infections, people of female sex with the CC genotype had a higher rate of spontaneous clearance of reinfection than other participants.^16^

HCV reinfections following cure with direct-acting antivirals (DAAs) may also clear spontaneously, although evidence remains limited. One recent study documented spontaneous clearance in one of 11 reinfections after successful DAA treatment, but eight had unknown outcomes, limiting estimation of the clearance probability.^19^ Therefore, an algorithm that separately accounts for primary infection, reinfection after spontaneous clearance, and reinfection after DAA cure can assign different probabilities to people with different clearance- related characteristics.

This variation in clearance across different subgroups is especially relevant to controlled human infection (CHI) models proposed for HCV vaccine development. In initial CHI studies, participants would receive a defined dose of virus and undergo scheduled follow-up to characterize the challenge virus and estimate the expected spontaneous clearance rate.^20^

These studies would inform the timing of treatment and the design of subsequent CHI vaccine trials.^20^ Differences in age, sex, or *IFNL4* genotype between initial CHI cohorts and groups subsequently enrolled in CHI vaccine trials could make it difficult to distinguish vaccine-induced clearance from spontaneous clearance. The expected small numbers of people recruited per group for any CHI model would make it difficult to stratify across all combinations of these factors. Although previous modeling studies stratify by sex,^4^ few quantify how outcomes vary across combinations of sex, *IFNL4* genotype, and age (**Table 1**). Such estimates could inform CHI model study design, where small sample sizes limit the ability to assess these combinations directly. Increasing the realism of simulations can address this gap in the existing literature by examining how participant characteristics impact spontaneous clearance, which may have utility in determining the optimal length of follow-up for a CHI model prior to a vaccine trial.^5, 20^

**Table 1.** Inputs and assumptions for the HCV clearance algorithm.

| Pathway | Input | Symbol | Value (95% CI) | Usage | Section | Source |
|---|---|---|---|---|---|---|
| Genotype assignment | CC probability: Hispanic, non-Hispanic Black, non-Hispanic White, Other | Pr(CC<sub>i</sub>) | 0.40, 0.16, 0.47, 0.86 | Assign CC genotype at initialization | **2.2** | Chen et al., 2024<sup>21</sup>; gnomAD v4.1.1<sup>22</sup> |
| First infection | Overall clearance | — | 0.26 (0.22–0.29) | Target mean used to determine intercept (*α*) | **2.3** (**Eq. 1**) | Micallef et al., 2006<sup>23</sup> |
| First infection | Female vs. male | *β*<sub>female</sub> | RR: 1.61 (1.42–1.83) | Risk ratio used to set the sex coefficient | **2.3** (**Eq. 1**) | Levinsson et al., 2024<sup>9</sup> |
| First infection | Age ≥45 vs. <45 years | *β*<sub>age</sub> | OR: 0.79 (0.64–0.97) | Age coefficient, log(*OR*) | **2.3** (**Eq. 1**) | Aisyah et al., 2018<sup>7</sup> |
| First infection | CC vs. CT/TT | *β*<sub>CC</sub> | OR: 2.75 (2.23–3.38) | Genotype coefficient, log(*OR*) | **2.3** (**Eq. 1**) | Miri et al., 2021<sup>8</sup> |
| First-infection timing | Cleared by 3, 6, 12, 24 months | — | 19.8%, 27.9%, 36.1%, 37.1%, cumulative | Rescale to 53.4%, 75.2%, 97.3%, 100.0% among selected clearers | **2.5.1** | Aisyah et al., 2018<sup>7</sup> |
| Reinfection after spontaneous clearance | Overall clearance by 180 days | — | 0.52 (0.33–0.73) | Target weighted mean used to set baseline cumulative hazard (*H*<sub>0</sub>) | **2.4.1** (**Eq. 2**) | Sacks-Davis et al., 2015<sup>16</sup> |
| Reinfection after spontaneous clearance | Female vs. male | *HR*<sub>i</sub> | HR: 2.45 (0.72–8.29) | Assigned to female non-CC agents | **2.4.1** (**Eq. 2**) | Sacks-Davis et al., 2015<sup>16</sup> |
| Reinfection after spontaneous clearance | Age >25 vs. ≤25 years | *HR*<sub>i</sub> | HR: 1.72 (0.45–6.53) | Age multiplier for >25 in every sex–genotype group | **2.4.1** (**Eq. 2**) | Sacks-Davis et al., 2015<sup>16</sup> |
| Reinfection after spontaneous clearance | CC vs. non-CC | *HR*<sub>i</sub> | HR: 2.00 (0.56–7.18) | Assigned to male CC agents | **2.4.1** (**Eq. 2**) | Sacks-Davis et al., 2015<sup>16</sup> |
| Reinfection after spontaneous clearance | Female CC vs. all other groups | *HR*<sub>i</sub> | HR: 4.16 (1.24–13.94) | Assigned to female CC agents | **2.4.1** (**Eq. 2**) | Sacks-Davis et al., 2015<sup>16</sup> |
| Timing after spontaneous clearance | Median clearance time | — | 90 days (IQR 60–132) | Log-normal, log SD 0.584, restrict to ≤180 days | **2.5.2** | Sacks-Davis et al., 2015<sup>16</sup> |
| Reinfection after DAA cure | Clearance by 1 year | *p*<sub>postDAA</sub> | 0.25, fixed | Primary-infection proxy, no covariate effects | **2.4.2** | Grebely et al., 2014<sup>15</sup> |
| Post-DAA timing | Cleared by 3, 6, 12 months among eventual primary clearers | — | 34%, 67%, 83%, cumulative | Rescale to 41.0%, 80.7%, 100.0% among one-year clearers | **2.5.3** | Grebely et al., 2014<sup>15</sup> |
| First or post-DAA infection without clearance | Mean time to chronic infection | — | 102 days, fixed | Exponential distribution | **2.5.1**, **2.5.3** | HepCEP timing rule<sup>4, 5</sup> |

Overall probabilities and the female-to-male risk ratio are values that the equations must reproduce as opposed to coefficient values themselves. Pr(CC<sub>i</sub>) is the probability of assigning CC genotype, and *p*<sub>postDAA</sub> is the fixed post-DAA probability. Timing values are cumulative, and the algorithm interpolates between their rescaled values. The reported reinfection median and IQR specify the underlying log-normal distribution before truncation. Abbreviations: CI = confidence interval, DAA = direct-acting antiviral, HR = hazard ratio, IQR = interquartile range, OR = odds ratio, RR = risk ratio, SD = standard deviation.

Our objective was to develop a modular, literature-informed algorithm of spontaneous HCV clearance for use in such simulations. We separately represented first infection, reinfection after spontaneous clearance, and reinfection after DAA cure. We incorporated age, sex, and *IFNL4* rs12979860 genotype and represented uncertainty in published estimates across runs. We used the agent-based model HepCEP (Hepatitis C Elimination in PWID) as a testing environment to examine whether the algorithm reproduced the intended person-specific probabilities and run- level variation. The present study tests the clearance calculation algorithm, and application to CHI model design is a proposed use of its modular structure.

## 2. Methods

### 2.1. Algorithm overview and evidence sources

Our clearance algorithm uses a simulated person’s demographic characteristics and infection and treatment history to assign a probability of spontaneous clearance. It then draws an individual outcome and, separately, a time to clearance or chronic infection (**Sections 2.3**–**2.5**). We tested the algorithm in HepCEP, an established ABM of HCV transmission among PWID.^4–6^ HepCEP supplies the population and transmission setting for this evaluation.

The algorithm follows three pathways: first infection, reinfection after spontaneous clearance, and reinfection after DAA cure (**Figure 1**). An agent with no previous HCV infection enters the first-infection pathway that uses age, sex, and *IFNL4* rs12979860 genotype to predict clearance. For reinfection, the pathway depends on whether the most recent infection cleared spontaneously or was cured with DAAs. This separation allows spontaneous clearance and treatment-induced cure to be associated with different subsequent clearance probabilities.

**Figure 1.**
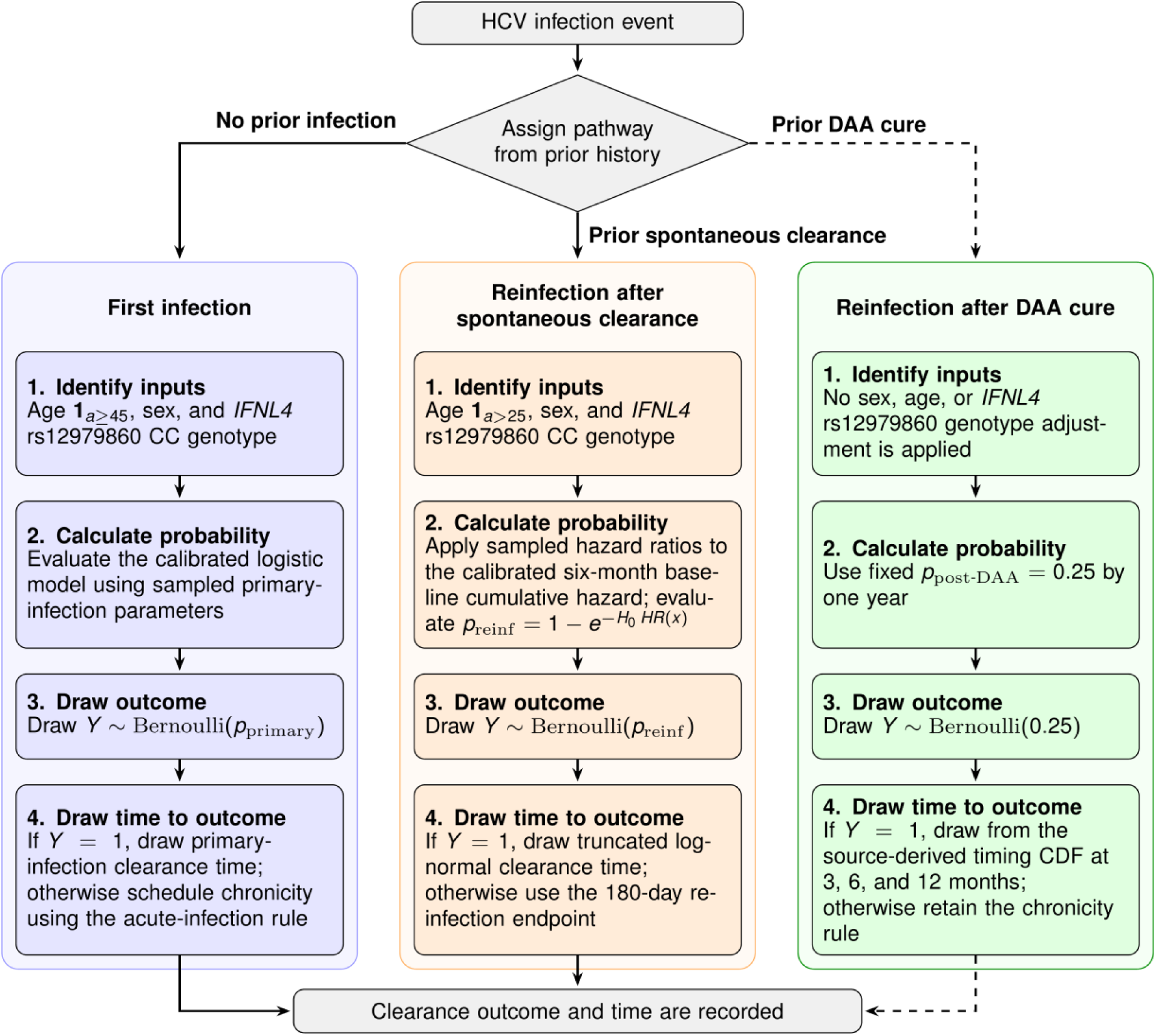
Pathways in the spontaneous-clearance algorithm. At each infection, the algorithm uses the person’s most recent clearance history to select first infection, reinfection after spontaneous clearance, or reinfection after DAA cure. It assigns a clearance outcome and a time to clearance or chronic infection. Dashed arrows mark the post-DAA pathway as one modeling assumption. An alternative could return DAA-cured individuals to the first-infection pathway. Reinfection pathways can be omitted in a simulation restricted to one exposure.

Reinfections after spontaneous clearance use age and combined sex–genotype groups to predict clearance,^16^ while reinfections after DAA cure use a fixed probability of 0.25 (**Section 2.4.2**).^15^

We used published estimates to describe clearance probabilities in our simulations, prioritizing systematic reviews and meta-analyses. For reinfection, we used the InC^3^ analysis, which combined data from several cohorts (**Table 1**).^16^ Estimates were selected for their relevance to spontaneous clearance among PWID, their compatibility with age, sex, genotype, and infection history, and the availability of uncertainty bounds. Where joint individual-level evidence was unavailable, we combined estimates from different studies and stated the assumptions involved. **Table 1** lists the parameter inputs, their sources, and the Equation and Methods subsection in which each is used.

### 2.2. *IFNL4* genotype assignment

We consider the C and T alleles of rs12979860 at the *IFNL4* locus (GRCh38, NC_000019.10:g.39248147C>T), where the CC (C/C) genotype is associated with a higher spontaneous clearance rate than the CT (C/T) or TT (T/T) genotypes.^8^ Our algorithm assigns each agent a CC or non-CC (CT/TT) genotype group at initialization. Individual genetic data were unavailable for the HepCEP test population, so assignment used probabilities for the agent’s race and/or ethnicity within HepCEP’s pre-existing categories: Hispanic, non-Hispanic Black, non-Hispanic White, or Other.^6^ The assigned CC probabilities were 0.40, 0.16, 0.47, and 0.86, respectively (**Table 1**).^21, 22^ These are observed rs12979860 CC genotype frequencies in gnomAD v4.1.1 genomes for admixed American, African/African American, and non-Finnish European populations. The value for the heterogeneous “Other” category used an East Asian population estimate. We chose the East Asian estimate because its C-allele frequency of 0.93 is the highest among gnomAD genetic ancestry groups. Together with the African/African American group, which has the lowest frequency (0.40), the four mapped groups reflect the full range of allele-frequency diversity across populations. Race and ethnicity only inform genotype assignment and do not enter the clearance equation as separate terms.

### 2.3. Clearance after first infection

For a first infection, the probability of spontaneous clearance depends on age group, sex, and CC genotype (**Table 1**). The female-to-male effect is reported as a risk ratio, which cannot be used in a logistic equation as an odds ratio. We therefore solve for the sex coefficient together with the intercept so that the population mean and the female-to-male risk ratio match their published estimates (**Table 1**).^9, 23^ **Equation 1** combines these terms with the age and genotype effects.

**Equation 1**. Probability of spontaneous clearance after first infection

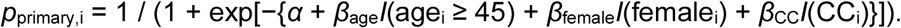

Each indicator *I*(·) equals 1 when its condition is true and 0 otherwise. At the literature point estimates, the age coefficient is *β*_age_=log(0.79), from the odds ratio for age ≥45 versus <45 years,^7^ and the genotype coefficient is *β*_CC_=log(2.75), from the odds ratio for CC versus CT/TT.^8^ The intercept *α* is chosen to match an overall clearance probability of 0.26,^23^ and *β*_female_ is chosen to match a standardized female-to-male risk ratio of 1.61.^9^ Each run uses sampled values of these estimates (**Section 2.6** and **Table 1**). Standard bisection searches, which repeatedly halve an interval containing the solution,^24^ determine the two unknown coefficients. Implementation details are in **Supplementary Methods S1**.

Published estimates carry uncertainty, so we allow model inputs to vary between simulation runs. Therefore, the same person can have a higher clearance probability in one simulation run than in another. However, within an individual run, two people with the same age, sex, and CC genotype receive the same clearance probability. In an individual run, a separate random draw determines each person’s outcome, so one person may still clear while the other develops persistent infection (**Section 2.6**).

### 2.4. Clearance after reinfection

#### 2.4.1. Reinfection after spontaneous clearance

For reinfection after spontaneous clearance, the algorithm matches an average clearance probability of 0.52 by 180 days, based on the InC^3^ Study.^16^ We used InC^3^ as the sole source of reinfection-clearance estimates because it pooled individual-level data from nine prospective cohorts and studied predominantly people who inject drugs, making its population relevant to application in HepCEP. It also provided both six-month cumulative clearance after reinfection estimates and reinfection-specific hazard ratios by age, sex, and CC genotype, including a combined sex–genotype contrast (**Table 1**).^16^ **Equation 2** converts these into subgroup probabilities using a proportional-hazards approximation.

**Equation 2**. Probability of clearance after reinfection following spontaneous clearance

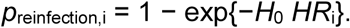

Here, *H*_0_ is a baseline cumulative hazard over 180 days and *HR*_i_ is the hazard-ratio combination for an agent. Sacks-Davis et al. reported separate comparisons adjusted for cohort site: age >25 vs. ≤25 years (HR: 1.72), female vs. male (HR: 2.45), CC vs. CT/TT genotype (HR: 2.00), and female CC versus all other participants (HR: 4.16).^16^ These were not jointly estimated subgroup effects, so as a modeling approximation, we assign multipliers of 1.00, 2.00, 2.45, and 4.16 to male non-CC, male CC, female non-CC, and female CC agents, respectively. We then multiply each sex–genotype group’s value by 1.72 for agents aged >25 years (**Table 1**).

The two age groups, two sex categories, and two *IFNL4* genotype groups produce eight combinations. We weight these combinations by their summed first-infection clearance probabilities, so the weights approximate the composition of people who have cleared a first infection and could enter this pathway. We choose *H*_0_ so that the weighted mean probability equals the sampled overall reinfection-clearance estimate. The bisection calculation is given in **Supplementary Methods S2**.

#### 2.4.2. Reinfection after DAA cure

Age, sex, and *IFNL4* genotype affect clearance of earlier infections modeled during simulation and can therefore influence which agents subsequently receive DAA treatment. However, chronic infections present at initialization are assigned using population data as opposed to the clearance algorithm. These infections accounted for an average of 91.2% of first DAA treatments across the 100 simulation runs. Age, sex, and genotype do not modify the post-DAA reinfection clearance probability because available studies have few reinfections, infrequent testing, or unknown outcomes, limiting the clearance estimates that can be transferred to the algorithm.^19, 25^ Therefore, we use a fixed probability of 0.25 by one year from a pooled analysis of nine prospective cohorts of primary acute infection (**Table 1**).^15^ This assumption can be revised as evidence accumulates (**Section 4.2**).

### 2.5. Time to clearance or chronic infection

The algorithm first draws whether an infection clears and then draws the time to that outcome.

Table 1 summarizes the timing inputs for each pathway.

#### 2.5.1. First infection

Each agent’s probability of spontaneous clearance after first infection is determined by **Equation 1**. For infections selected to clear, we then assign a clearance time. The published cumulative clearance estimates are 19.8%, 27.9%, 36.1%, and 37.1% by 3, 6, 12, and 24 months, respectively.^7^ To describe timing of HCV clearance, we divide each estimate by the 24- month estimate of 37.1%. Thus, among infections selected to clear, 53.4% clear by 3 months (19.8/37.1), 75.2% by 6 months (27.9/37.1), 97.3% by 12 months (36.1/37.1), and 100% by 24 months (37.1/37.1). Just over half therefore clear within three months, and only approximately 2.7% clear between 12 and 24 months. The algorithm draws each clearance time by linearly interpolating between these cumulative values.

For first infections that do not clear spontaneously, the algorithm draws a time to transition to the chronic-infection state from an exponential distribution with a mean of 102 days, using the established HepCEP timing rule (Table 1).^4, 5^ This mean applies only to infections pre-selected to not clear.

#### 2.5.2. Reinfection after spontaneous clearance

The InC^3^ study reported a median clearance time of 3.0 months with an interquartile range of 2.0–4.4 months for individuals reinfected after spontaneous clearance.^16^ We use these data to specify a log-normal distribution with an underlying median of 90 days and a log-scale standard deviation of 0.584, then restrict draws to ≤180 days (Table 1). The restriction is a modeling choice and changes the distribution of accepted times. Reinfections that do not clear are classified as chronic at 180 days.

#### 2.5.3. Reinfection after DAA cure

For reinfection after DAA cure, we use Grebely et al.’s primary-infection data to model both the probability of clearance and its timing (**Section 2.4.2**).^15^ Among participants who cleared during follow-up, 34%, 67%, and 83% had cleared by 3, 6, and 12 months.^15^ We restrict this timing distribution to clearance within one year and similarly divide each percentage by 83%. Among post-DAA reinfections selected to clear, the resulting cumulative percentages are therefore 41.0% by 3 months (34/83), 80.7% by 6 months (67/83), and 100% by 12 months (83/83) (Table 1). Times are drawn by linear interpolation between these cumulative values. Post-DAA reinfections that do not clear use an exponential time to chronic infection with a mean of 102 days.^4, 5^

### 2.6. Parameter uncertainty

In the first infection and reinfection after spontaneous clearance pathways, published estimates have uncertainty. At the start of a run, the algorithm draws one value for each uncertain input using the point estimate and its reported 95% confidence interval (Table 1). Overall probabilities are sampled from normal distributions on the logit scale, while risk ratios, odds ratios, and hazard ratios are sampled from normal distributions on the log scale. We then transform the draws back to the original scales, and the coefficients and baseline hazard are chosen to match that run’s sampled inputs. All agents share these inputs within the run, and a new run receives a new set. For example, one run may use a higher overall clearance estimate than another, while agents within either run still have different chances according to their characteristics.

This variation across runs represents uncertainty in the published evidence. The separate random draw for each infection represents whether that individual infection clears. Literature inputs are sampled independently because their joint uncertainty was not reported. The post- DAA probability and timing parameters are fixed, so uncertainty is not applied in this manner.

### 2.7. Testing the clearance algorithm

We evaluated the algorithm in a fixed reference cohort (**Figure 2**) and in full HepCEP transmission simulations. The reference cohort contained 8,000 agents. Calculations at literature point estimates and across 10,000 parameter draws isolated subgroup probabilities and uncertainty without running transmission.

**Figure 2.**
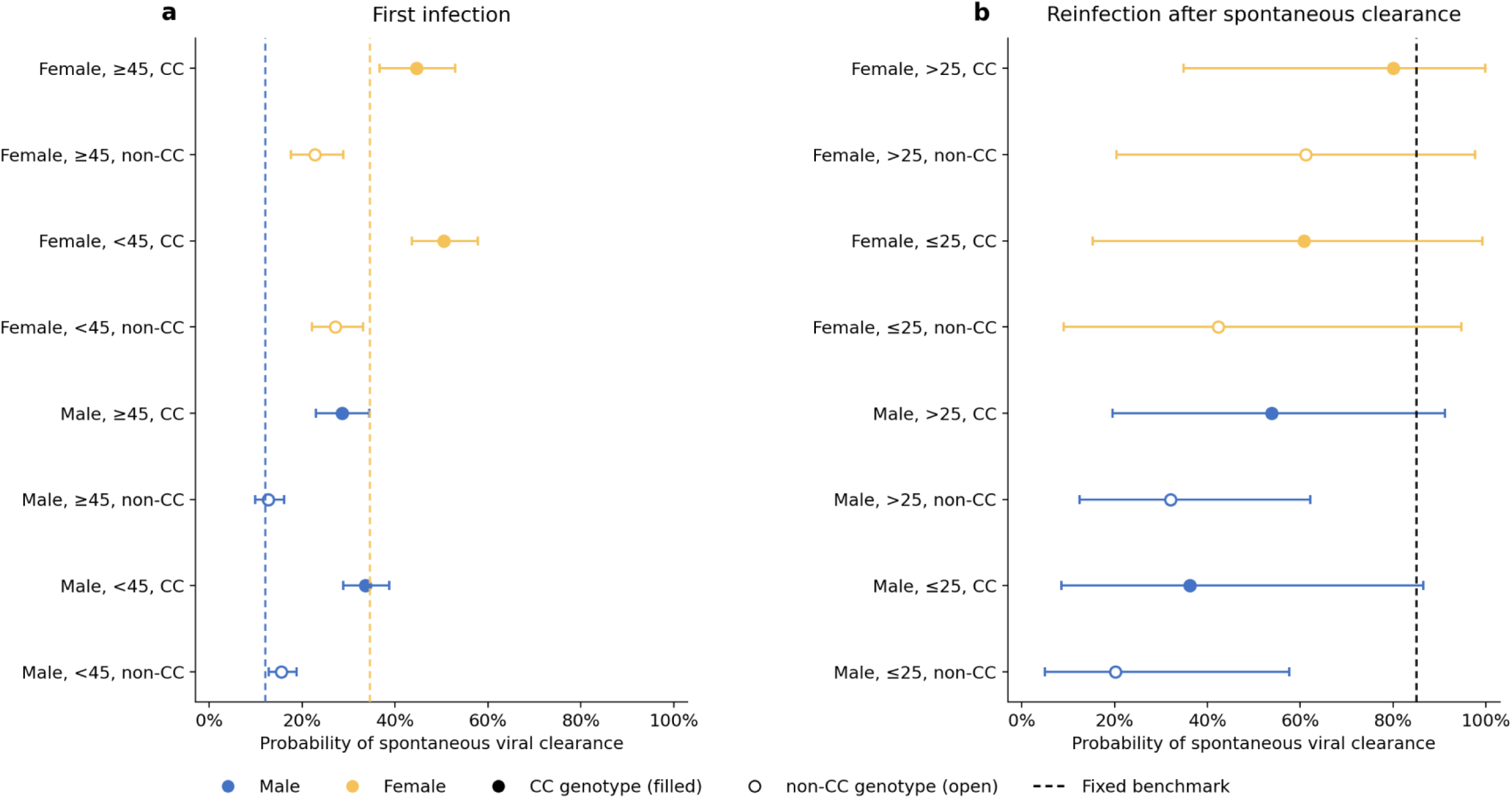
Subgroup clearance probabilities and parameter uncertainty. Panels show first infection (**a**) and reinfection after spontaneous clearance (**b**) in a fixed reference cohort of 8,000 agents without running transmission. The algorithm uses literature point estimates, and error bars show central 95% intervals across 10,000 parameter draws. These intervals describe uncertainty in input estimates as opposed to individual infection outcomes. Fixed benchmark probabilities are included as reference values, and reinfection after DAA cure is not depicted here, as it was represented with a fixed clearance probability of 0.25.

For the full simulations, we ran 100 matched-seed pairs (seeds 42–141). Each pair contained one run using our clearance algorithm and one using a fixed-probability benchmark drawn from established HepCEP settings.^4–6^ This benchmark provides a reference for the numerical evaluation. Both runs used an 800-day horizon, an initial population of 32,000 agents, and the same demographic, network, treatment, burn-in, and other non-clearance settings wherever applicable. Every algorithm run sampled one literature-parameter set. Assigned probabilities and outcomes were recorded at the clearance-decision point without an additional random draw. For infections acute at initialization, clearance decisions are recorded immediately by the algorithm and at acute resolution by the benchmark. The benchmark values are reported with the results in **Section 3.3.**

Within each run, pathway, and subgroup, probabilities were placed in 30 common bins from 0 to 1 and normalized to sum to one. Agents were grouped by sex and genotype while pooling ages, or by sex and age while pooling genotypes. Bin masses were averaged across the 100 runs, giving each run equal weight. We also calculated the number of recorded clearance decisions and the mean assigned probability within each run, then summarized their means and standard deviations across runs. These summaries include all recorded decisions.

### 2.8. Software and data

The clearance algorithm was implemented in Python and tested within HepCEP. Analysis scripts retain pathway labels, assigned probabilities, sampled parameters, outcomes, and timing so that calculations can be checked at both run and agent levels. This study used published summary estimates and simulated agents and did not involve human participants or identifiable data.

## 3. Results

### 3.1. First-infection clearance probabilities

At the literature point estimates, first-infection clearance probabilities ranged from 0.505 for female CC agents aged <45 years to 0.127 for male non-CC agents aged ≥45 years (**Figure 2a**). Among female agents aged <45 years, the clearance probability was 0.271 without CC genotypes and 0.505 with CC genotypes. The corresponding male clearance probabilities were 0.155 and 0.336. For age ≥45 years, these values were 0.227 and 0.446 in female agents and 0.127 and 0.285 in male agents, respectively. The central 95% interval was 0.436–0.578 for female CC agents aged <45 years and 0.099–0.161 for male non-CC agents aged ≥45 years (**Figure 2a**). Although some intervals overlapped, point estimates were consistently higher with CC genotype and female sex and lower in the older age group.

### 3.2. Reinfection clearance probabilities

After spontaneous clearance, reinfection probabilities covered a wider range (**Figure 2b**). For agents aged ≤25 years, point-estimate probabilities were 0.201 for male non-CC, 0.362 for male CC, 0.424 for female non-CC, and 0.608 for female CC agents. The age >25 hazard ratio increased these to 0.321, 0.539, 0.612, and 0.800, respectively. Parameter-uncertainty intervals were narrower for first infection than for reinfection after spontaneous clearance. Central 95% intervals extended from 0.050–0.577 in male non-CC agents aged ≤25 years to 0.349–0.998 in female CC agents aged >25 years.

The post-DAA pathway assigned a reinfection clearance probability of 0.25 to every subgroup. No uncertainty interval is shown because this input is fixed.

### 3.3. Testing in full transmission simulations

The fixed-probability benchmark assigned first-infection clearance probabilities of 0.121 for male agents and 0.346 for female agents (**Figure 3a**) and a clearance probability of 0.85 for reinfection after spontaneous clearance (**Figure 3d**).^4, 5^ Our benchmark implementation fixes spontaneous clearance following reinfection after DAA cure to zero, differing from the post- treatment behavior described in the published HepCEP model.^6^ At literature point estimates, differences between algorithm first-infection probabilities and these reference values ranged from −11.9 percentage points for female non-CC agents aged ≥45 years to +21.5 percentage points for male CC agents aged <45 years. All eight reinfection point estimates were below the benchmark value of 0.85, reflecting the literature mean of 0.52 used by the algorithm.

**Figure 3.**
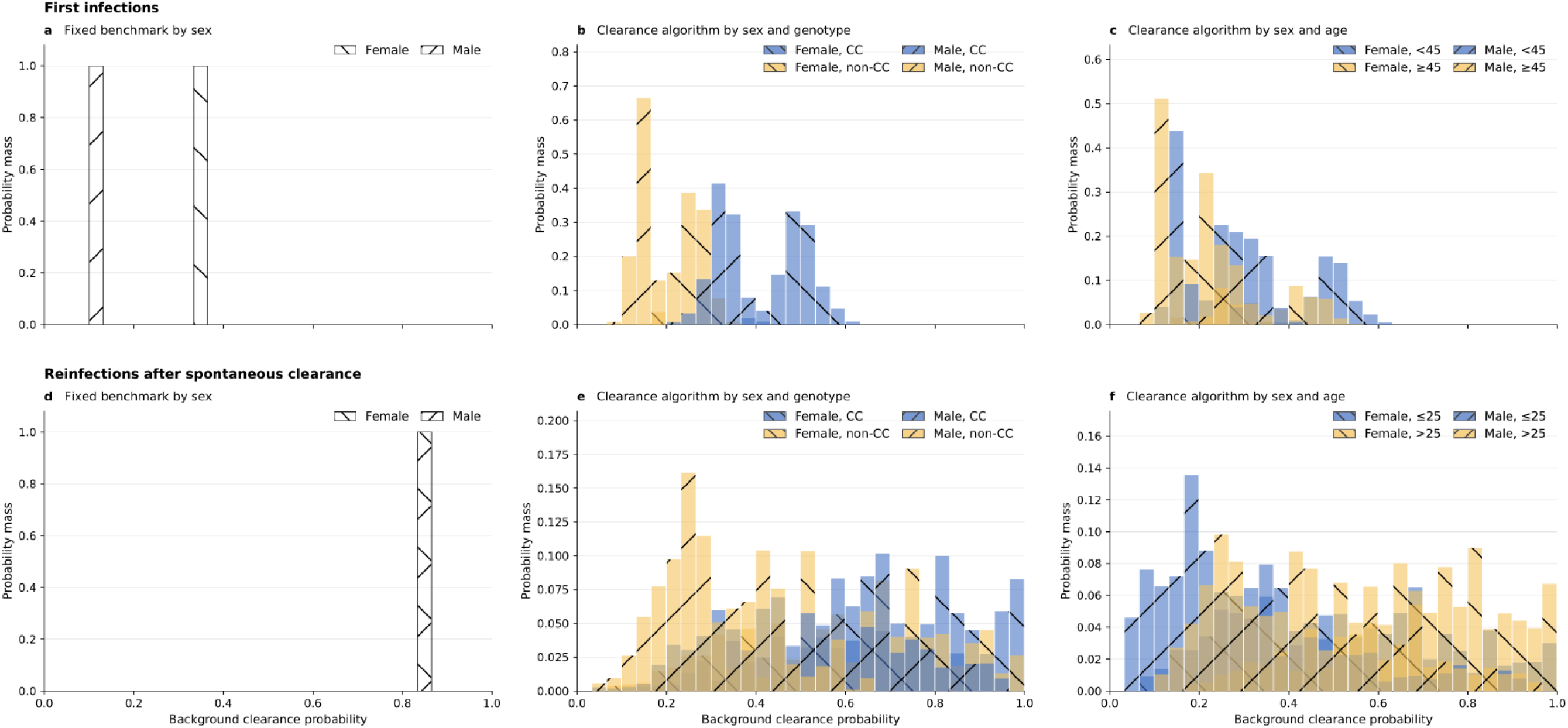
Assigned background clearance probabilities in transmission simulations. Histograms show the fixed benchmark for first infection by sex (**a**), the algorithm for first infection by sex and *IFNL4* rs12979860 genotype with ages pooled (**b**), and by sex and age (<45 vs. ≥45 years) with genotypes pooled (**c**). Reinfection after spontaneous clearance is shown for the benchmark by sex (**d**), and for the algorithm by sex and genotype with ages pooled (**e**) and by sex and age (≤25 vs. >25 years) with genotypes pooled (**f**). Female groups precede male groups in each legend. Panels **a** and **d** have unfilled bars with black outlines. Blue and yellow denote CC and non-CC in **b** and **e**, and younger and older ages in **c** and **f**. Left-leaning hatching denotes female and right-leaning hatching denotes male. Each run and subgroup is normalized over 30 common bins from 0 to 1 before bin masses are averaged across 100 runs. Each run therefore receives equal weight. Reinfection after DAA cure is not depicted here, as it was represented with a fixed clearance probability of 0.25.

All 200 simulations in the 100 matched-seed pairs completed. Algorithm runs recorded a mean of 1,063.25 clearance decisions for first infection (SD 36.99), 498.50 for reinfection after spontaneous clearance (48.79), and 215.01 for post-DAA reinfection (16.39) per run.

Corresponding benchmark counts were 973.15 (38.00), 595.44 (47.70), and 160.46 (12.86) (Table 2).

**Table 2.** Assigned clearance probabilities and recorded decision counts in 100 matched-seed pairs of transmission simulations.

| **Infection pathway** | **Clearance rule** | **Recorded per-run clearance decisions** | **Mean assigned clearance probability** |
|---|---|---|---|
| First infection | Fixed benchmark | 973.15 (38.00) | 0.200 (0.004) |
| First infection | Clearance algorithm | 1,063.25 (36.99) | 0.270 (0.019) |
| Reinfection after spontaneous clearance | Fixed benchmark | 595.44 (47.70) | 0.850 (fixed) |
| Reinfection after spontaneous clearance | Clearance algorithm | 498.50 (48.79) | 0.523 (0.109) |
| Reinfection after DAA cure | Fixed benchmark | 160.46 (12.86) | 0 (fixed) |
| Reinfection after DAA cure | Clearance algorithm | 215.01 (16.39) | 0.250 (fixed) |

Values are mean (SD) across 100 runs unless fixed. Mean probabilities are calculated within each run and then summarized across runs, giving each run equal weight. Abbreviations: DAA = direct-acting antiviral, SD = standard deviation.

For first infection, the algorithm distributed assigned probabilities across age, sex, genotype, and run-level parameter draws (**Figures 3b** and **3c**). The mean across runs was 0.270 (SD 0.019). With equal weight assigned to each run, the episode-level probability distribution had a median of 0.262 and an interquartile range of 0.155–0.339. Benchmark probabilities were concentrated at 0.121 and 0.346 (**Figure 3a**), with a mean of 0.200 (0.004), a median of 0.121, and an interquartile range of 0.121–0.346.

For reinfection after spontaneous clearance, the algorithm generated a broad distribution reflecting covariate differences and uncertainty across runs (**Figures 3e** and **3f**). Its mean assigned probability was 0.523 (SD 0.109), the equal-run-weighted median was 0.515, and the interquartile range was 0.324–0.700. The benchmark probability was fixed at 0.850 (**Figure 3d**). In the post-DAA pathway, clearance probabilities were fixed at 0.25 for the algorithm and zero for the benchmark (Table 2).

## 4. Discussion

### 4.1. Principal findings and applications

We developed an algorithm of spontaneous HCV clearance that accounts for age, sex, *IFNL4* rs12979860 genotype, and three infection histories. Within each run, the algorithm matches the overall probabilities from published estimates, and agents with varying characteristics have different chances of clearance. Testing in 100 HepCEP runs (paired with 100 fixed-probability benchmark simulations) reproduced the intended differences between people and between runs. An infection selected to clear may still be treated, or follow-up may end, before its scheduled clearance time. Among first infections selected to clear spontaneously, 53.4% are scheduled to clear within three months and 75.2% within six months, but only 2.7% are scheduled to clear between 12 and 24 months. For reinfection after DAA cure, 80.7% of infections selected to clear are scheduled to do so within six months and the remaining 19.3% between six and 12 months. Under our first-infection timing distribution, approximately 24.8% of infections selected to clear spontaneously would not yet have cleared at six months. We recently simulated 9 months of follow-up in HCV vaccine efficacy trials to determine whether HCV infection became chronic or cleared spontaneously.^5^ Further simulations are needed to determine whether shorter follow-up, with DAA treatment beginning before 9 months after infection, would allow this distinction in CHI models. Earlier treatment could prevent observation of later spontaneous clearance, potentially affecting estimates of vaccine efficacy.

First-infection probabilities varied almost fourfold, from 0.127 to 0.505 across the eight subgroups. This variation supports accounting for age, sex, and *IFNL4* genotype when assigning clearance probabilities in individual-based simulations. For reinfection after spontaneous clearance, the InC^3^ estimates describe clearance rates over 180 days and include a combined female–CC genotype effect.^16^ **Equation 2** retains these features and yields probabilities that can be used in individual-based simulations. Male, non-CC agents had low probabilities of clearing a first infection, and if reinfected after spontaneous clearance, they also had low probabilities of clearing again (**Figure 2**). Male agents aged ≤25 years had lower reinfection-clearance point estimates than those aged >25 years with the same genotype (**Figure 2b**), reflecting the age hazard ratio used in **Equation 2**. Its wide confidence interval and the wider reinfection intervals reflect the uncertainty of these estimates.

Realistic clearance probabilities could support CHI model simulations. For a study with one controlled exposure in HCV-naïve participants, the first-infection calculation could use participants’ measured genotypes and recruitment characteristics. Small CHI cohorts might preferentially recruit participants with the favorable *IFNL4* CC genotype to reduce the risk of persistent infection. This would increase expected spontaneous clearance and could make comparisons with later cohorts misleading if their genotype composition differs. Varying participant composition and clearance assumptions could help assess sample size and interpret clearance at the planned endpoint.^20^ These applications remain to be tested.

A next step is to assess how much spontaneous clearance would occur before treatment. Combining our first-infection probabilities and timing distribution gives estimated six-month clearance probabilities of 9.5%–38.0% across the eight host subgroups at the literature point estimates, assuming no treatment before six months. The current model uses the same timing distribution among first infections selected to clear, and estimating subgroup-specific clearance times would require additional data. These estimates could inform the follow-up period before treatment in initial CHI studies. Earlier DAA treatment would shorten the observation period and prevent later spontaneous clearance from being observed.^20^ Future CHI model simulations could use more detailed timing data to estimate how many clearances would be missed and how earlier treatment might affect DAA duration and costs.

Notably, some of our timing parameters were also derived from chimpanzee studies and may not generalize to humans.^4^ Viral-kinetic profiles based on human data include clearance courses extending beyond six months after primary infection and reinfection.^26^ Our algorithm could also be coupled to HepCEP implementations that represent viral kinetics. In a recent HepCEP study, Tatara et al. (2026) incorporated individual viral-load trajectories during acute infection, reinfection, and DAA treatment, with transmission probabilities depending on viral load.^26^ Our algorithm could supply the probability of entering a clearing or persistent course, while a viral-kinetics module supplies the trajectory. Such coupling would require profile selection and timing consistent with the assigned outcome. This approach has not been tested here, but it could connect literature-informed clearance heterogeneity to simulations that also represent the changing risk of HCV transmission for an infected individual.

### 4.2. Limitations

Published estimates were combined across studies with different populations, acute-infection definitions, follow-up schedules, and levels of adjustment. Matching the reported averages aligns the algorithm’s clearance probabilities with the literature estimates, but it does not fully account for confounding or other differences between populations. Estimates largely derived from people who inject drugs may not transfer directly to healthy volunteers in CHI models. In the absence of empirical HCV CHI clearance data, this work uses estimates from naturally acquired infection to inform the timing of DAA treatment initiation in future human CHI studies.

The present algorithm assumes HCV genotype 1. This is the most prevalent viral genotype in the United States,^27^ and much of the *IFNL4* evidence used here concerns genotype 1.^8, 11–14^ Application to other viral genotypes requires appropriate estimates and validation. In particular, the host-genotype effect should not be assumed to capture differences between the viruses used in CHI model studies.

Age is represented by two categories, with different cut points for first infection and reinfection, following the published estimates. *IFNL4* genotype assignment in the HepCEP test population uses HepCEP’s broad race and ethnicity categories that do not capture all allele-frequency variation. The “Other” category is especially uncertain, and gnomAD v4.1.1 genetic ancestry groups do not correspond exactly to HepCEP’s race and ethnicity categories. Measured genotypes would avoid these assignment assumptions.

Literature inputs are sampled independently because joint uncertainty was unavailable. The reinfection algorithm combines separately estimated contrasts into joint subgroup multipliers, although these joint effects were not estimated in the original study.^16^ Timing parameters are fixed, and the first-infection timing curve is approximated by rescaling pooled cumulative estimates from a different source than the overall clearance estimate. Truncating the reinfection timing distribution at 180 days also changes its median and quartiles. The post-DAA probability and timing use primary-infection evidence as opposed to evidence specific to reinfection after DAA cure. Finally, we did not evaluate CHI model sample size, vaccine efficacy estimates, elimination targets, or treatment costs.

## 5. Conclusion

Our literature-informed algorithm represents spontaneous HCV clearance by age, sex, *IFNL4* rs12979860 genotype, and infection history. It also accounts for uncertainty in published parameter estimates and variation in whether an individual infection clears. Testing in HepCEP reproduced the intended subgroup and run-level variation and preserved literature-derived population means. Its modular structure allows reinfection pathways to be disabled for future single-exposure CHI model simulations and supports refinement of the post-DAA assumptions. External validation and study-specific scenarios are needed before using it to guide CHI model design or other intervention decisions.

## Artificial Intelligence Usage

OpenAI and Anthropic models (GPT-6.1 Sol and Claude Opus 5.5, respectively) were used to assist the development of code for this work and preparation of the manuscript. The authors take full responsibility for this work.

## Data Availability

Code and simulation data will be deposited on GitHub and Zenodo upon journal publication. In the interim, all code and data are available from the corresponding authors upon request.

