## Supplementary Methods for "Modeling spontaneous hepatitis C virus clearance by age, sex, *IFNL4* genotype, and infection history: Implications for simulations of elimination and vaccine trials"

### S1. Solving the first-infection equation

**Equation 1** contains two unknowns: the intercept $\alpha$ and the female coefficient $\beta_{\text{female}}$. Their values must reproduce both the overall clearance probability and the standardized female-to-male risk ratio (**Section 2.3** and **Table 1**). We use nested bisection: a standard interval-halving search for one unknown inside the search for the other.[1]

After the population is initialized and a run's literature inputs are sampled, the outer search proposes a value of $\beta_{\text{female}}$. For each proposal, an inner search varies $\alpha$ over [−20, 20] until the mean **Equation 1** probability across the starting population's age, sex, and genotype composition matches the sampled overall probability (point estimate 0.26).[2] This wide numerical bracket is on the log-odds scale, so for an intercept alone, its endpoints correspond to probabilities of approximately $2 \times 10^{-9}$ and $1 - (2 \times 10^{-9})$. It provides a broad search range as opposed to a plausible biological interval for clearance.

Holding age and genotype fixed, we next calculate a mean probability after assigning every agent female sex and a mean after assigning every agent male sex. The outer search varies $\beta_{\text{female}}$ over [−20, 20], repeating the inner search at every step, until the ratio of these standardized means matches the sampled risk ratio (point estimate 1.61).[3] Each search performs 80 iterations and retains the final bracket midpoint. The resulting coefficients reproduce the two sampled inputs to numerical precision while retaining the sampled age and genotype odds ratios. This step is necessary because the logarithm of a risk ratio is not the corresponding logistic coefficient.

### S2. Solving the reinfection equation

The starting population is divided into eight categories defined by two age groups (≤25 and >25 years), two sex categories, and two genotype groups (CC and non-CC), as described in **Section 2.4.1**. For each category, we sum the agents' **Equation 1** probabilities and normalize the eight sums to obtain weights $w_k$ that total one. Categories more likely to clear a first infection therefore receive greater weight in the pool eligible for reinfection after spontaneous clearance.

For candidate values of $H_0$, the solver evaluates

$$\sum_{k=1}^{8} w_k[1 - \exp(-H_0\, HR_k)]$$

using the hazard-ratio combinations in **Table 1**. The lower bracket is zero and the initial upper bracket is one. The upper bracket is doubled until the calculated mean reaches the sampled overall reinfection-clearance probability (point estimate 0.52).[4] Bisection then halves this bracket

80 times and retains its midpoint.[1] The weighted mean 180-day probability consequently matches the sampled overall input while retaining the model-assigned subgroup hazard multipliers.
